# DISTRIBUTION AND GENOMIC CHARACTERIZATION OF COLOMBIAN CARBAPENEM-RESISTANT *Acinetobacter baumannii* ISOLATES

**DOI:** 10.1101/2025.08.25.25334258

**Authors:** Diego Prada-Cardozo, Jaime Moreno, Verónica Rincón, Sandra Yamile Saavedra, María T. Reguero, Carolina Duarte, Jhon Donato, Camila Pérez, Laurent Falquet, Emiliano Barreto-Hernández

**Author notes:** Corresponding autor Grupo de Microbiología, Instituto Nacional de Salud, Avenida Calle 26 No 51-20, Bogotá, Colombia.

## Abstract

**Objectives:** To characterize carbapenem-resistant *Acinetobacter baumannii* isolates associated with healthcare-associated infections (HAIs) in Colombia using whole-genome sequencing (WGS).

**Methods:** A total of 83 *A. baumannii* isolates collected between 2012 and 2015 were sequenced using the Illumina platform. Raw reads were assembled by SPAdes and classified via multilocus sequence typing (MLST), using the Pasteur, Oxford, and core genome MLST (cgMLST) schemes. Antimicrobial resistance determinants, capsule types, and outer core (OC) serotypes were identified. Phylogenetic relationships were inferred through single nucleotide polymorphism (SNP) analysis using IQ-TREE.

**Results:** MLST analysis assigned isolates to 10 sequence types (STs) in the Pasteur scheme and 20 STs in the Oxford scheme. The most prevalent types were ST79^P^ (68.7%) and ST124^°^ (25.3%). IC5 (ST79^P^) and IC7 (ST25^P^/ST945^P^) were the predominant ICs, confirming their endemic presence in Colombia and highlighting the genetic diversity of emerging clonal variants. cgMLST identified 13 minimum spanning trees (MSTSs), with MST1 grouping 19 isolates. Carbapenem resistance was primarily driven by the presence of the class D β-lactamase gene *bla*OXA-23. The most common K locus types were KL9 and KL151, while the dominant OC locus was OCL10. Phylogenetic analysis revealed two major clades and seven minor clusters.

**Conclusions:** This study provides a comprehensive genomic overview of carbapenem-resistant *A. baumannii* isolates circulating in Colombia, highlighting key resistance determinants and population structure. These findings can inform public health strategies to mitigate the spread of high-risk clones and improve antimicrobial resistance surveillance.

## 1. Introduction

Carbapenem-resistant *Acinetobacter baumannii* (CRAB) is a major cause of healthcare-associated infections (HAIs) worldwide, including ventilator-associated pneumonia, wound infections, and bloodstream infections, these infections are particularly challenging to manage due to limited therapeutic options (1). CRAB develops resistance through diverse molecular mechanisms, including horizontal gene transfer mediated by plasmids and mobile genetic elements, which facilitate the dissemination of antibiotic resistance genes. The accumulation of several resistance genes makes this microorganism a potential multidrug-resistant (MDR) bacterium (2).

Population structure analysis of *A. baumannii* has contributed to understanding its dynamics and behavior in clinical environments. These studies have been based mainly on different genotyping methods: multilocus sequence typing (MLST), which includes two schemes, Oxford (3) and Pasteur (4), and has become the reference method (5), and whole-genome sequencing (WGS), which at present has been used to characterize bacterial strains with high resolution compared to other techniques (6). Worldwide, *A. baumannii* has shown the presence of international clones (IC) 1, 2, and 3, of which clone 2 and its divergent strains have spread more frequently than the other two ICs (4). However, other minor clones have been reported (7).

Epidemiological studies in Colombia indicate a rising prevalence of antimicrobial resistance in *A. baumannii,* highlighting its growing clinical significance and impact on healthcare settings (8). In 2005, 2008, 2009, and 2014, *A. baumannii* was reported to carry resistance to carbapenems in approximately 38%, 41%, 45.5%, and 56% of the isolates, respectively (9–11). The most common carbapenem resistance mechanisms found in these microorganisms are carbapenemases, represented mainly by class D (OXA-23-like, OXA-24-like, and OXA-58-like) and to a lesser extent by class B (NDM, VIM, IMP) (12–14).

This study aimed to analyze the genomic determinants responsible for antimicrobial resistance profiles, the epidemiology and phylogenetic relationship of 83 CRAB from 14 Colombian states (46 hospitals) from 2012–2015.

## 2. Materials and methods

### 2.1. Description of the isolates, identification, and antimicrobial susceptibility

Between 2012 to 2015, 188 carbapenem-resistant *Acinetobacter* spp. isolates (14 in 2012, 42 in 2013, 45 in 2014, and 87 in 2015) were sent to the Microbiology Group of the National Reference Laboratory (NRL) of the Instituto Nacional de Salud (INS) for phenotypic and genotypic characterization as part of voluntary, laboratory-based surveillance of antimicrobial resistance in HAIs. Isolates were reidentified up to the *Acinetobacter baumannii-calcoaceticus* complex using Vitek-2 compact® (bioMérieux SA, Marcy-l’Etoile, France) and/or Phoenix^TM^ (Becton Dickinson). Antimicrobial susceptibility profiles for imipenem (IMP), meropenem (MEM), ceftazidime (CAZ), cefepime (FEP), ampicillin-sulbactam (SAM), ciprofloxacin (CIP), gentamicin (GEN), amikacin (AMK) and piperacillin-tazobactam (PTZ) were determined by the disk diffusion (DD) method and interpreted according to the parameters of the Clinical & Laboratory Standards Institute (15). MDR isolates were those nonsusceptible to at least one agent in three or more antimicrobial categories. A subset of 83 CRAB isolates, collected from 46 hospitals across 14 departments (political states), was selected for WGS at the molecular epidemiology and bioinformatics laboratories of the National University of Colombia. The selection was based on the following criteria: molecular confirmation of *A. baumannii* via *rpoB* amplification by PCR; isolates associated with HAIs; year of isolation between 2012 and 2015; and carbapenem resistance determined by DD.

### 2.2. DNA extraction, sequencing, and assembly

*A. baumannii* isolates were cultured in Luria Bertani broth and incubated at 37 °C until obtaining an optical density of 0.6 to 0.8 (λ 600 nm). DNA extraction was performed following the instructions of the UltraClean^®^ Microbial DNA Isolation Kit (MOBIO Laboratories, Inc.). A NanoDrop® 2000 spectrophotometer (Thermo Scientific™) and agarose gel (0.8%) were used to evaluate the concentration and purity of DNA. Purified bacterial DNA was sent to Macrogen, Korea, for WGS by the HiSeq 2000 platform (Illumina, San Diego, CA). Library preparation was performed using the Illumina TruSeq Nano kit. Paired-end reads of 101 bp in length were produced, the quality was analyzed with FastQC v0.12.1 (16), and read cleaning was performed with Trimmomatic v0.39 (17). Genomes were assembled by the *de novo* method Spades v.3.8 (18), and genome characteristics and quality were obtained by QUAST v5.2.0 (19).

### 2.3. Genomic identification, typing and identification of resistance genes

Molecular identification and typing were performed using MLST 1.8 with the Pasteur and Oxford schemes (20). Novel alleles and sequence types (ST) were submitted to the MLST database. Lipooligosaccharide outer core loci (OCL), capsular polysaccharide loci (KL), and biofilm-related genes were identified by Pathogenwatch v12.0.4 https://pathogen.watch/, and PubMLST (20). Additionally, cgMLST was determined by Ridom SeqSphere+ software v.8.2.0 (21), and a clonal relationship analysis was carried out based on 2390 target genes of strain ACICU by the scheme for *A. baumannii* (22). The distances between their allelic profiles were visualized in a minimum spanning network using the parameter ‘pairwise ignoring missing values’. Antibiotic resistance mediator genes and mutations were identified by the ABRicate program v1.0.1 (23) https://github.com/tseemann/abricate, in which the following databases were included: NCBI (24), CARD (25), ARG-ANNOT (26) Resfinder (27), MEGARES (28), and PlasmidFinder (29). The K and OC locus were identified by the Pathogenwatch platform v12.0.4 https://pathogen.watch/.

### 2.4. Comparative genomics and phylogeny

Single nucleotide polymorphism (SNP) calling was performed by Snippy v2 (30). IQ-TREE software v2.3.3 (31) was used to infer the phylogenetic relationship, including *A. baumannii* ATCC 19606 as a reference genome (GenBank accession number **CP045110.1**). Finally, the phylogenetic tree was edited with FigTree v1.4.4 (32) and visualized with Microreact (33).

## 3. Results

### 3.1. Origin of the isolates

Eighty-three *A. baumannii* isolates collected between 2012 and 2015, but mainly in 2014 (n=29, 34.9%) and 2013 (n= 26, 31.3%), were sequenced. Isolates came from 14 Colombian departments, of which Valle del Cauca (n=14, 16.9%), Antioquia (n=14, 16.9%) and Santander (n=12, 14.4%) were the primary sources. Isolates were obtained from several tissues and fluids, mainly from skin and soft tissues (n=27, 32.5%) and upper and lower respiratory tract (n=21, 25.3%). Hospitalized patients were mainly old-aged adult (age > 45 years; n=49, 59%) and male patients (n=55, 66.3%) (Table 1).

**Table 1.** Origin of the isolates.

|  | Origin of the Isolate | No. of Isolates | % |
| --- | --- | --- | --- |
| **Year of Isolate** | 2012 | 7 | 8.4 |
|  | 2013 | 26 | 31.3 |
|  | 2014 | 29 | 34.9 |
|  | 2015 | 21 | 25.3 |
| **Age Groups** | Child (0-16) | 3 | 3.6 |
|  | Young Adult (17-30) | 12 | 14.4 |
|  | Middle-aged Adult (31-45) | 13 | 15.7 |
|  | Old-aged Adult (Above 45) | 49 | 59 |
|  | ND | 6 | 7.2 |
| **Sample of Isolate** | Skin and Soft Tissue | 27 | 32.5 |
|  | Upper and Lower Respiratory Tract | 21 | 25.3 |
|  | Bloodstream | 15 | 18.1 |
|  | Sterile Body Fluids | 8 | 9.6 |
|  | Urine | 8 | 9.6 |
|  | Bones | 3 | 3.6 |
|  | Gastrointestinal Tract | 1 | 1.2 |

ND: No data

### 3.2. Characteristics of genomes

The properties of WGS are summarized in Supplementary Table 1. The genome size for all sequenced isolates remained close to the expected size, with an average of 4.042.638 bp. The highest and lowest N50 values were 423.623 bp and 52.691 bp, respectively, with an average of 150.265 bp. The annotation variables showed an average CDS of 3.857, 63 tRNA, three rRNA, and one tmRNA. values within the expected ranges, according to the summary of the sequenced genomes in the NCBI (https://www.ncbi.nlm.nih.gov/genome/?term=Acinetobacter+baumannii+genome).

### 3.3. MLST typing

According to MLST typing, isolates were classified into 10 Pasteur (ST^P^) and 20 Oxford (ST^°^) (Table 2). Three ST^P^s (ST^P^920, ST^P^944 and ST^P^945) and nine ST^°^s (ST^°^2122, ST^°^2123, ST^°^2124, ST^°^2125, ST^°^2126, ST^°^2127, ST^°^2128, ST^°^2129, and ST^°^2131/ST^°^2140) were novel and reported in the pubMLST database (Table 2). ST79^P^ (n=57, 68.7%), ST25^P^ (n=14, 16.9%), ST124^°^ (n=21, 25.3%), and ST758^°^ (n=19, 22.9%) were the dominant STs according to the Pasteur and Oxford MLST schemes, respectively. Other less frequent ST^P^s accounted for 14.4% (n=12) and ST^°^s accounted for 51.8% (n=43) of the isolates.

**Table 2.**
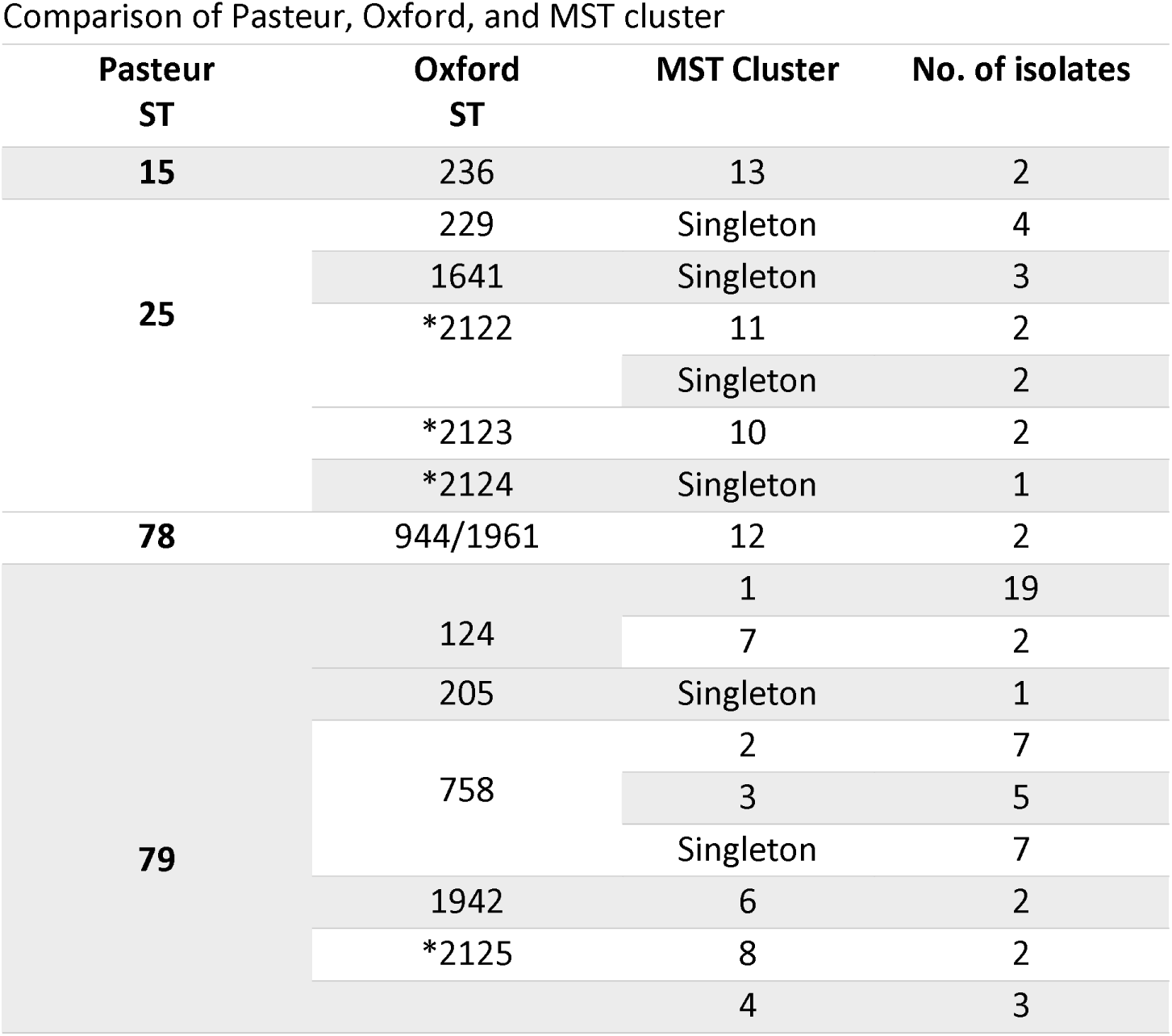
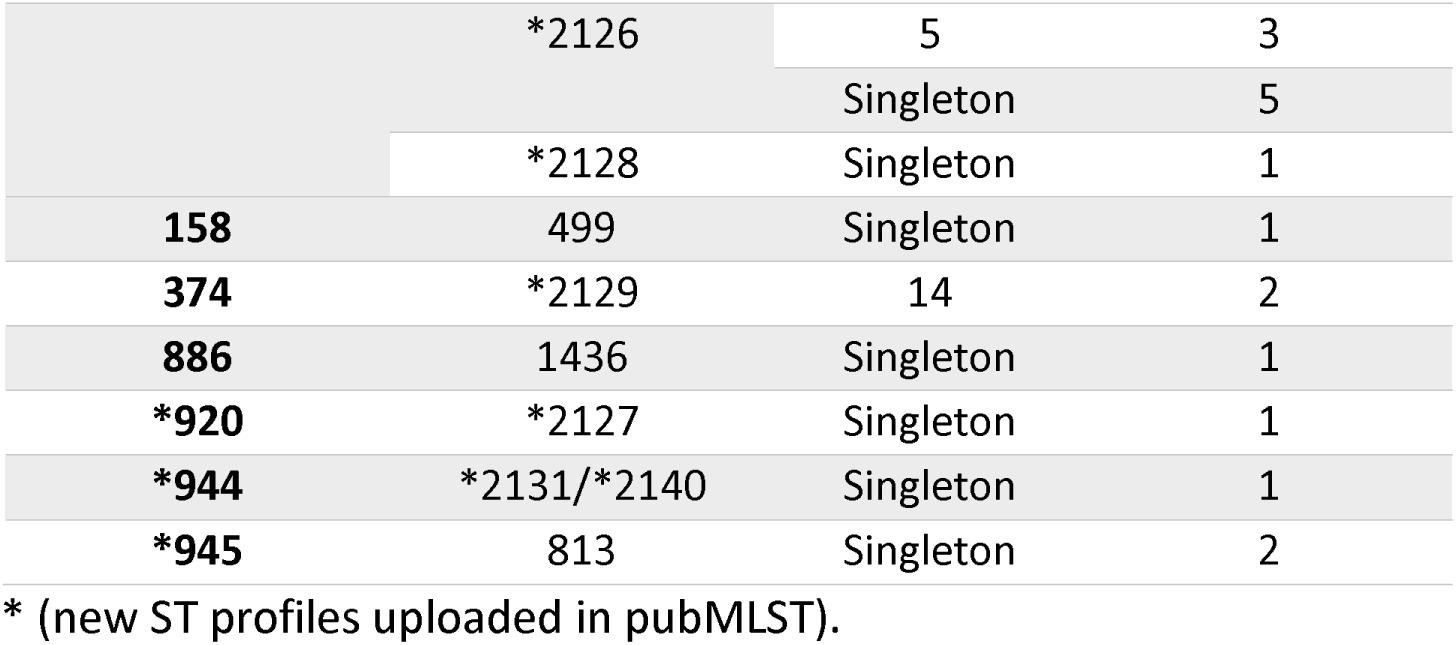
Comparison of Pasteur, Oxford, and MST cluster

### 3.4. cgMLST typing

cgMLST analysis identified 13 clusters or minimum spanning tree (MST), consisting of closely related genotypes (≤10 allele differences) and 30 isolates were assigned as singletons (Figure 1A). Seven of the 13 MST groups included isolates from the same location and year (MST-4, MST-7, MST-8, MST-9, MST-10, MST-12, MST-13), which means possible direct nosocomial transmissions since these groups are not found in other places, nor are they prolonged in time. Instead, the main group, MST-1, comprised 19 ST79^P^/ST124^°^ isolates from different departments, and all except one isolate were resistant to all the evaluated antimicrobials. According to the higher resolution of cgMLST, ST79^P^ isolates were divided into eight clusters and 14 singletons; moreover, the second most prevalent ST, ST25^P^, and its single locus variant (SLV) ST945^P^ showed two groups and 12 singletons, showing more significant differences between the alleles of these isolates than the ST79^P^ isolates.

**Figure 1.**
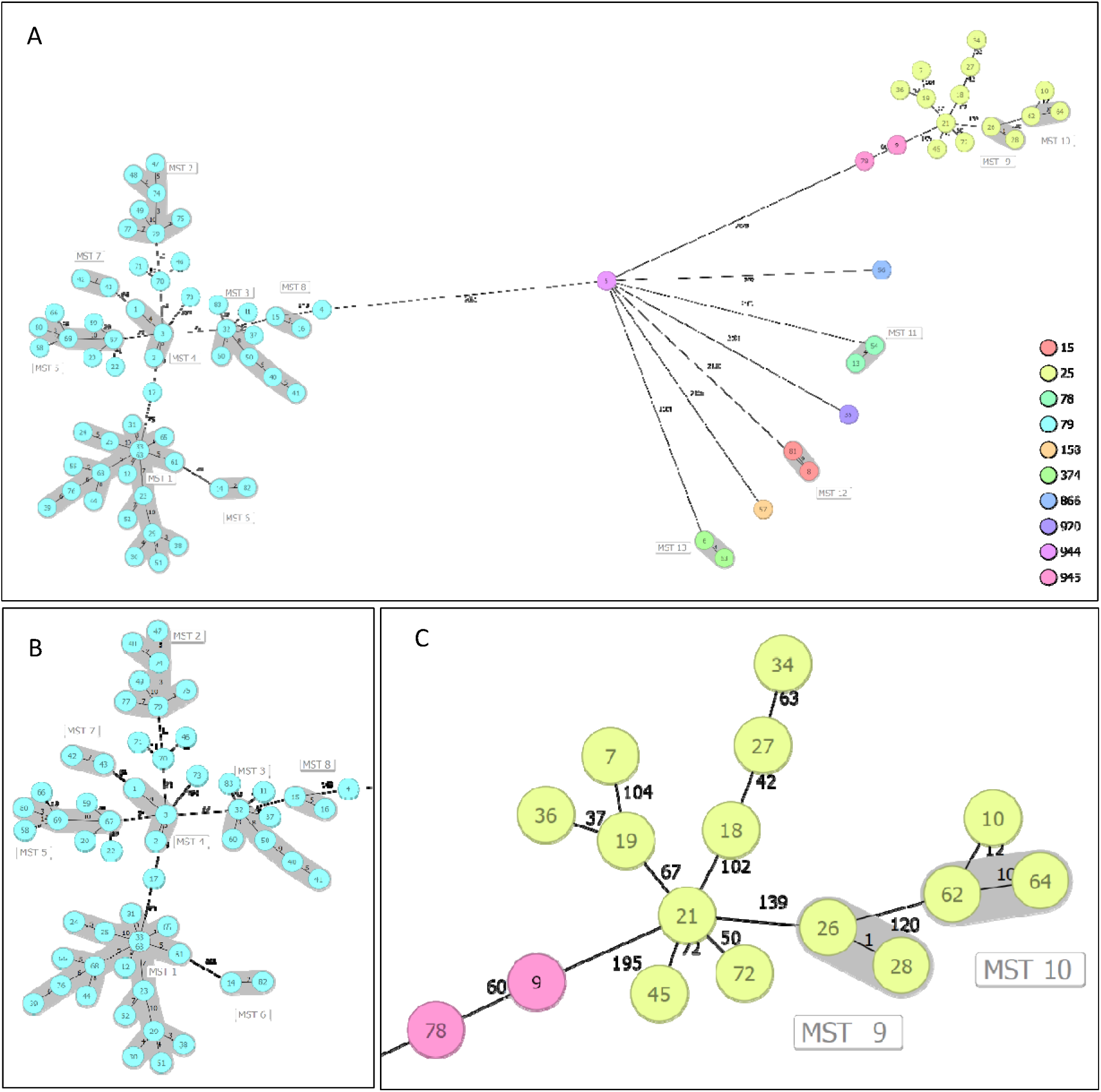
**A:** Minimum spanning tree of the 83 carbapenem-resistant *Acinetobacter baumannii* isolates based on core genome multilocus sequence typing (cgMLST). Nodes represent the isolates which are colored by ST Pasteur, MST clusters are shaded, and numbers between the nodes indicate the number of allelic differences. Major ICs are shown, IC5 (**B**) and IC7 (**C**).

### 3.5. Analysis by clades

The major clade of isolates was related to the IC5; this clade grouped 57 isolates (68.7%). These isolates were genetically very close, with maximum SNPs of 1959 and all the isolates belonged to ST79^P^, meanwhile by the Oxford scheme seven STs were identified, which ST124^°^ (n=21; 36.8%) and ST758^°^ (n=19; 33.3%) were the principal. Eight cgMLST (n=43; 75.4%) and 14 singletons were identified in this clade (Figure 1A, 1B). All the isolates, except one, were OCL10 serotype. Seven K locus were identified; K151 and K9 were associated with ST124^°^ and ST758^°^, respectively. The isolates of this clade came from eleven departments, of the four years of study, Figure 2. All isolates carried *bla*OXA-23, 49 isolates presented *bla*OXA-65, and the other 8 isolates presented *bla*OXA-64 and distinctively were ST2126^°^ and KL16; and recovered mainly from Antioquia, finally 82.4 % of the isolates carried TEM-1.

**Figure 2.**
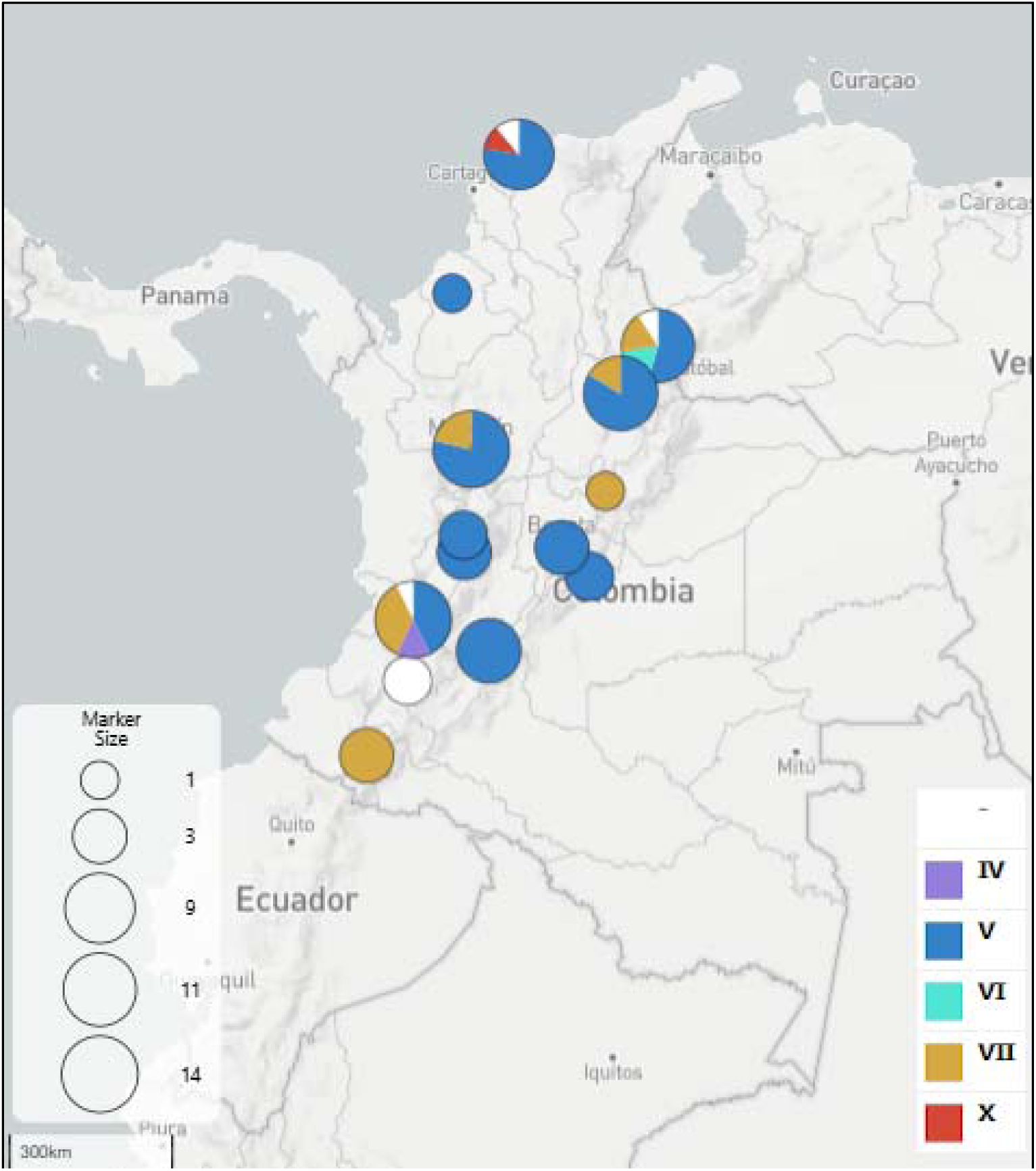
Distribution of the isolates by International Clones (IC). Five ICs were identified, other isolates were not identified. The size of each circle is proportional to the number of isolates recovered in each state.

The second largest clade was related to IC7; this clade grouped 16 isolates (19.3%), with maximum SNPs of 2627. Isolates belonged to ST25^P^ (n=14), and ST945^P^ (n=2), meanwhile by the Oxford scheme six STs were identified, which ST229^°^ (n=4; 25%) and ST2122^°^ (n=4; 25%) were the principal. Two cgMLST (n=4; 25%) and 12 singletons were identified in this clade (Figure 1A, 1C). Two OCL serotypes were identified: OCL5 (n=8; 50%) and OCL6 (n=8; 50%). Seven K locus were identified; K167 was the most prevalent (n=4; 25%) and was associated with ST2122^°^. The isolates of this clade came from six departments, of the four years of study, Figure 2. In general, this clade was more variable and presented more enzymes different from the OXA type: 13 isolates carried *bla*OXA-23, and all isolates carried *bla*OXA-64. Eight isolates had TEM-1, two carried NDM-1, and one carried KPC-2.

Clade IC4 was identified in two ST15^P^/ST236^°^ isolates with 267 SNPs. These isolates generated the CgMLST12 (Figure 1A), were related to OCL7 and K9 and came from the Valle del Cauca department in the year 2013, and carried *bla*OXA-23 and *bla*OXA-51, Figure 2, 3.

Clade IC6 was identified in two ST78^P^/ST944^°^ isolates with 292 SNPs. These isolates generated the CgMLST11 (Figure 1A). Isolates were related to OCL1 and K3 and came from the Norte de Santander department in 2013 and 2014 years and carried *bla*OXA-23, *bla*OXA-90, and CTX-M-115, Figure 2, 3.

Clade IC10 was represented just for one ST158^P^/ST499^°^ isolate. This isolate was singleton by cgMLST (Figure 1A) and was related to OCL1 and K6. The isolate came from Atlántico department in 2013 and carried *bla*OXA-23, *bla*OXA-65, and VIM-4, Figure 2, 3.

The other four clades were unrelated to any IC. One clade is represented in two ST374^P^/ST2129^°^ isolates with 177 SNPs. These isolates generated the CgMLST13 (Figure 1A). Isolates were related to OCL11 and K69 and came from the Cauca department in 2012, Figure 2. Isolates carried *bla*OXA-146 (OXA-23-like), *bla*OXA-90, and one had VIM-4. The other 3 clades are represented by one sample each. ST866^P^/ST1436^°^ isolate which was singleton, OCL2, K9, from Valle del Cauca in 2015 and carried *bla*OXA-385, NDM-1, TEM-1 and VEB-9. ST920^P^/ST2127^°^ isolate, singleton, OCL4, K172, from Norte de Santander in 2014 and carried *bla*OXA-106 (OXA-51-like), *bla*OXA-72 (OXA-24-like) and *bla*OXA-255 (OXA-143-like). ST944^P^/ST2131^°^ isolate which was singleton, OCL14, K52, from Atlántico in 2012 and carried *bla*OXA-23, *bla*OXA-120 and VIM-4, Figure 1A, 2.

### 3.6. Resistance

All isolates were resistant to IMP and PTZ, which were also resistant to FEP (98.8%), MEM (97.7%), CIP (95.3%), SAM (80%), CAZ (91.9%), AMK (91.9%) and GEN (88.4%) (Table 3). All isolates had OXA-51-like carbapenemase and *Acinetobacter*-derived cephalosporinase 2 (ADC-2), which are intrinsic to *A. baumannii*. OXA-23-like + OXA-51-like was identified in 78 isolates (94%), followed by NDM-1 + OXA-51-like (n=3; 3.6%), VIM-4 + OXA-23-like + OXA-51-like (n=3; 3.6%), KPC-2 + OXA-51-like (n=1; 1.2%), and OXA-24-like + OXA-143-like + OXA-51-like (n=1; 1.2%), (Figure 3, Supplementary Table 1).

**Table 3.**
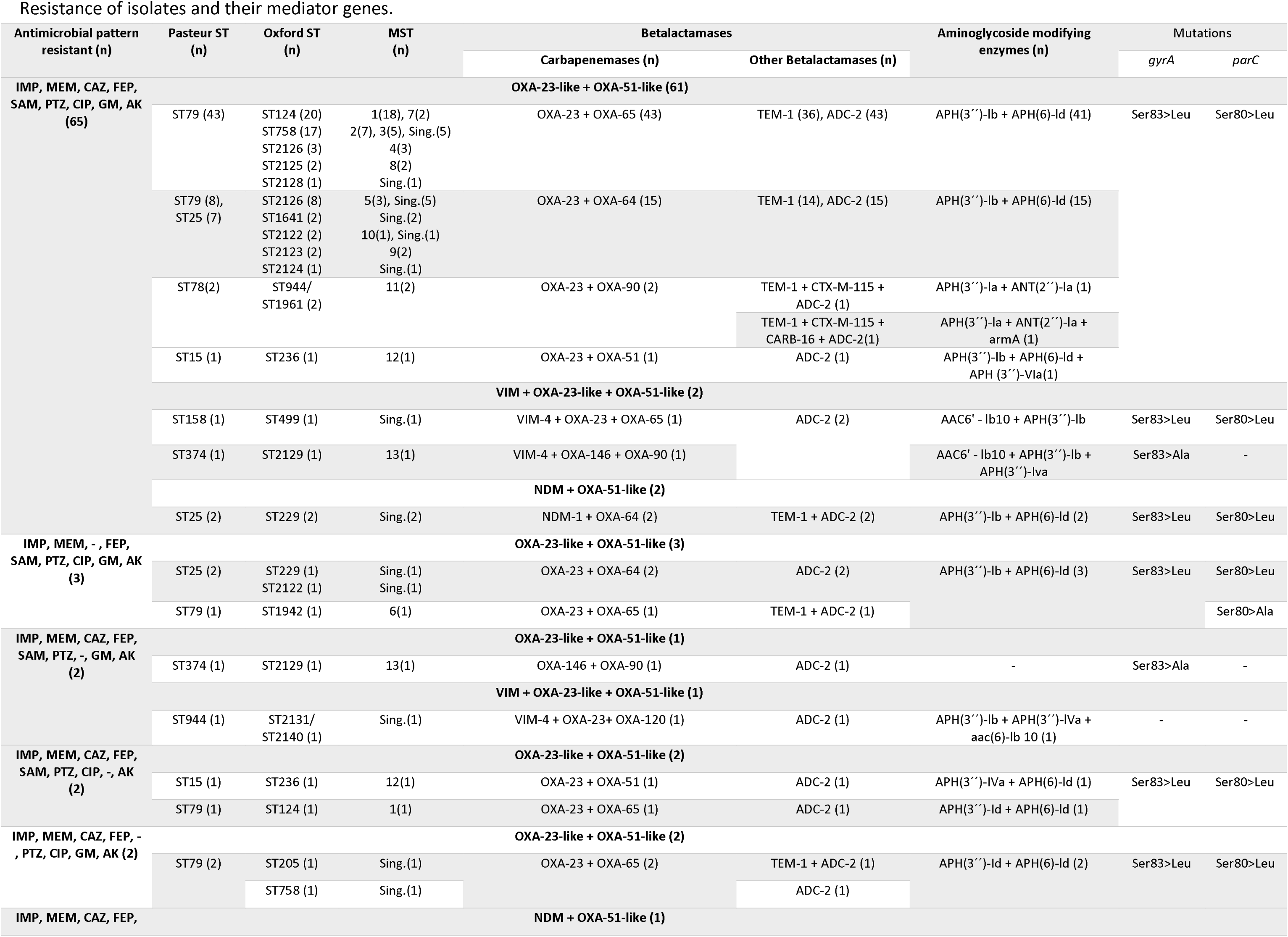
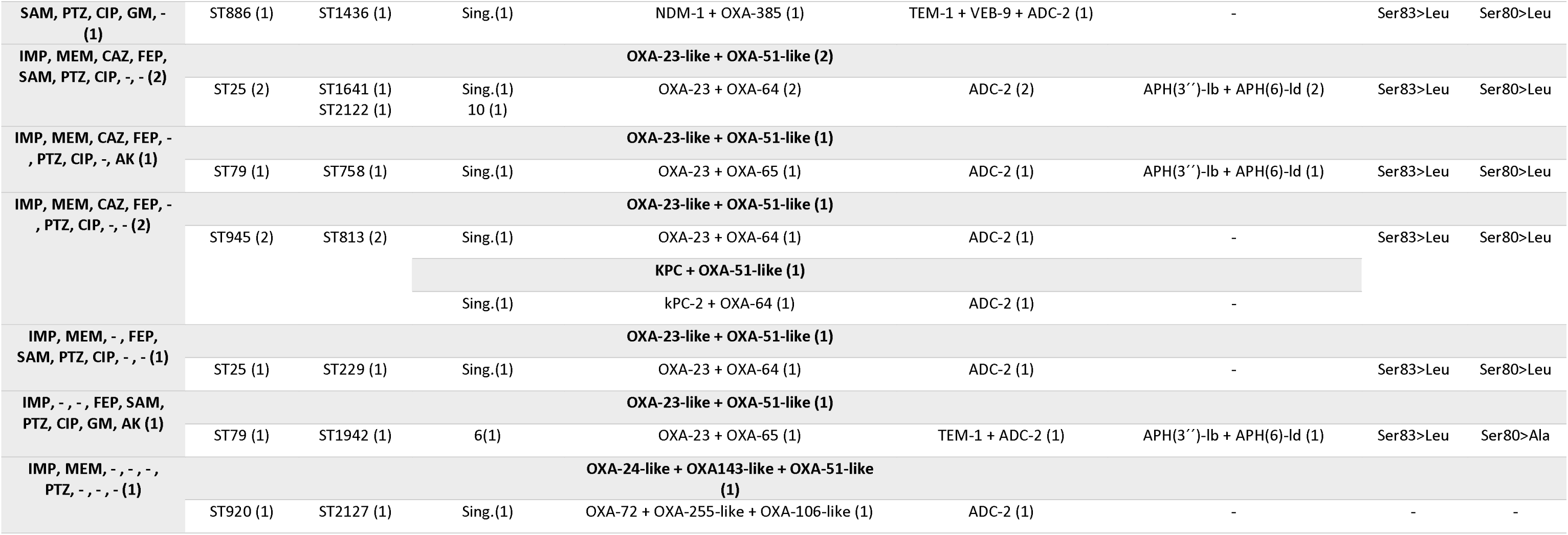
Resistance of isolates and their mediator genes.

**Figure 3.**
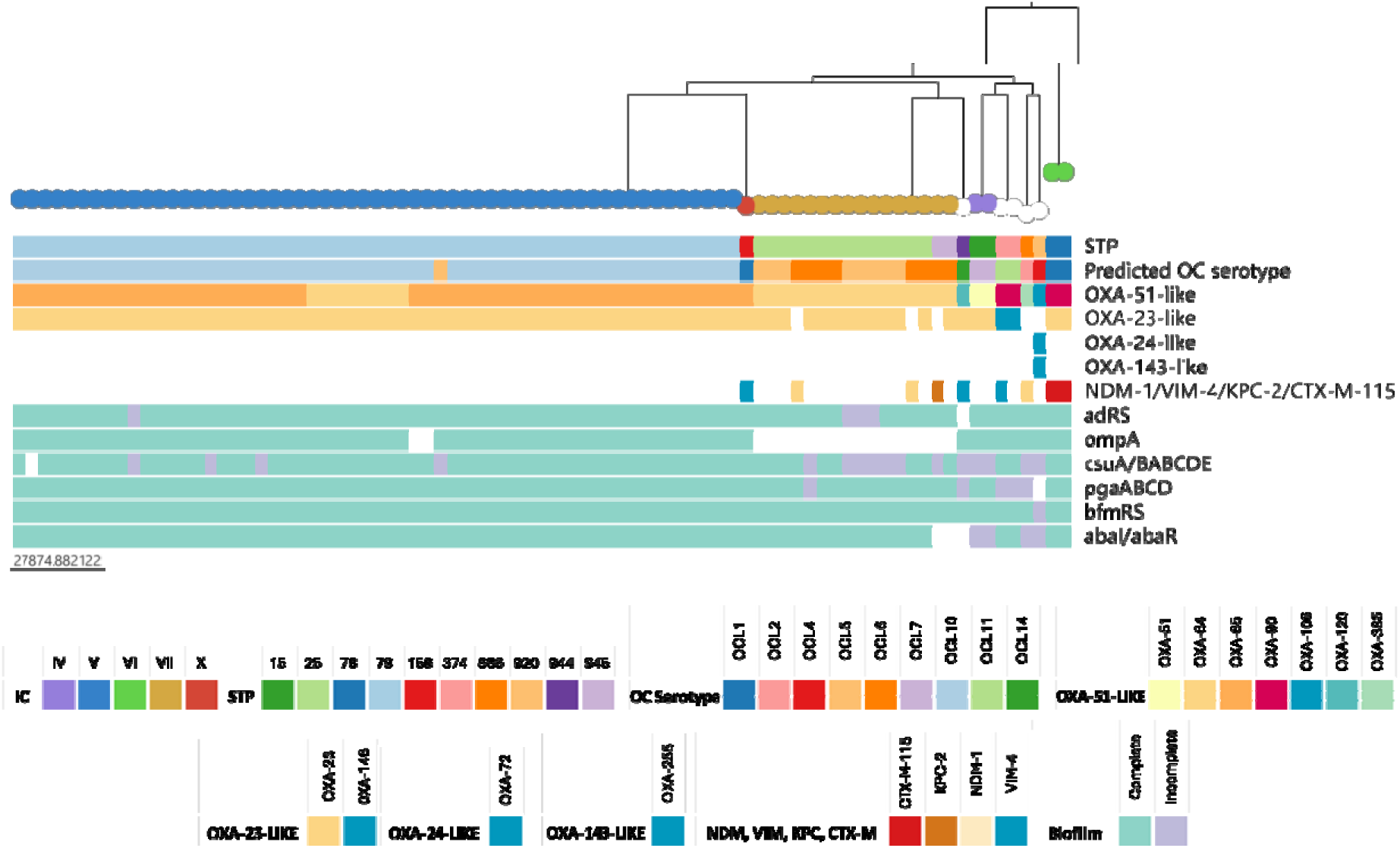
Phylogenetic relationship of the 83 isolates is represented on the top colored by International Clones. Metadata in blocks on the bottom represent in order: Pasteur ST, predicted OC serotype, carbapenemases and biofilm genes. https://microreact.org/project/cBr2FH4Mjge16gp7GopS64-acineto.

Aminoglycoside phosphotransferases APH(3’’)-Ib and APH(6)-Id were found in 86.7% (n=72) and 84.3% (n=70) of the isolates, respectively. Mutations encoding Ser83>Leu in the *gyrA* gene and Ser80>Leu in the *parC* gene were detected in 92.8% (n= 77) of the isolates. Additionally, two new mutations were identified: i) Ser83>Ala in the *gyrA* gene was found in two isolates (ST374^P^/ST2129^°^) without mutations in the *parC* gene, of which one was resistant to CIP, and the other was susceptible. ii) Ser80>Ala mutation in the *parC* gene was found in two quinolone-resistant isolates (ST79^P^/ST1942^°^) that also had Ser83>Leu mutation in the *gyrA* gene. On the other hand, 30 different genomic resistance profiles were identified in all the isolates, and the most frequent profile was found in 29 ST79^P^ isolates with 19 genetic elements associated with resistance (Supplementary Table 1).

### 3.7. Biofilm

Biofilm-related genes were identified, including *bfmRS* (two components system, which upregulates the csu pili operon) (n=82; 98.8%), incomplete *bfmRS* (n=1; 1.2%), *csuA/BABCDE* (chaperone-usher pili) (n=66; 79.5%), incomplete *csuA/BABCDE* (n=16; 19.3%), one isolate did not present the operon. *pgaABCD* (n=77; 92.8%), incomplete *pgaABCD* (n=5; 6%), one isolate did not present the operon. *ompA* (outer membrane protein) was identified in 65 isolates (78.3%). Isolates without *ompA* (n=18; 21.7%), interestingly, most of these (n=16; 88.9%) were related to IC7. Furthermore, quorum sensing genes *adRS* were found in 78 isolates (94%), incomplete form of *adRS* in four isolates (4.8%), *abaIR* (n=76; 91.6%) and incomplete *abaIR* (n=4; 4.8%).

## 4. Discussion

Antibiotic resistance in *A. baumannii* has been a growing concern since the 1970s, with a progressive increase in resistant strains worldwide (34). In 2018, resistance rates for imipenem (57.5%) and meropenem (48.5%) were reported in Colombian adult intensive care units (ICUs), highlighting the growing challenge of carbapenem-resistant *A. baumannii* infections (8), considered the last therapeutic option for MDR *A. baumannii* infections. In this study, we found *A. baumannii* isolates mainly related to IC5, ST79^P^, OCL 10, and OXA-23 as the principal carbapenemase, followed by isolates associated with IC7, which was more diverse genetically.

Most isolates came from soft tissues and upper and lower respiratory tract. These data agree with other studies that report nosocomial infections caused by *A. baumannii* associated with skin and soft tissue and pneumonia associated with mechanical ventilation and bloodstream infections (35–37). Patients most frequently affected by *A. baumannii* infections were old-aged adults (above 45) and middle-aged adults (31–45), probably because these patients may be more likely to be colonized and infected by resistant isolates due to previous exposures to antibiotics during their life (38).

We identified ten different Pasteur STs corresponding to 20 Oxford STs; this is because the genes of the Oxford scheme have more alleles than the Pasteur scheme. Although the two schemes are widely used, some studies have considered that the Pasteur scheme is more appropriate for population and epidemiological studies, and to compare *A. baumannii* with international isolates (39). On the other hand, analysis by cgMLST is more robust due to the comparison of allelic profiles, in this case, based on the 2390 target genes according to the scheme published by Higgins et al. (22).

Currently, CRAB isolates have been grouped into 11 ICs (40). Global analysis of CRAB found that IC5 was predominant in Latin America, and this lineage has a distinct emergence of clades with particular geographic dominance (41–44). For our study, IC5 (ST79^P^) was the most prevalent IC, which was characterized by the presence of OXA-23 as the main beta-lactamase in all CRABs. In addition, the isolates were found in 11 departments out of 14 sampled and in the 4 years of study, demonstrated that IC5 (ST79^P^) was widely present in Colombian territory and reinforces that it is endemic to Latin America (45,46), which suggests a potential regional adaptation or selective advantage of this lineage. Previous studies show that this IC has been circulating in Colombia since 2008 in Bogotá (12), persisting over time. The IC5 isolates have a close relationship despite having the greatest number of isolates compared with other ICs, presenting few differences at the level of SNPs compared to the other groups and representing a successful clonal group that has been established and spread in various regions of Colombia.

IC7 (ST25^P^ and ST945^P^) was the second predominant IC in this study; IC7 has been documented widely in Latin America, for example, in Brazil (47), Bolivia (48,49), Argentina (14), and Venezuela (43). In Colombia, IC7 circulated with isolates classified as ST229^°^ from Medellin city (37). The IC7 isolates had OXA-64, the characteristic *bla*OXA-51 variant from this lineage. Both, IC5 and IC7 are currently recognized as other lineages that contribute to the global distribution and dispersal of CRAB associated with the *bla*OXA-23 gene (50,51). IC7 was more diverse than IC5 and the genetic diversity is mainly due to homologous recombination, which can significantly affect phenotypic diversity (52). The identification of the new STs, ST920^P^, ST944^P^, and especially the ST945^P^ within the IC7 and its SLV of ST25^P^ may indicate the possible emergence of new lineages in Colombia. The emergence and proliferation of new clonal lineages in *A. baumannii* is due to high genomic plasticity, characterized by recombination and horizontal gene transfer (52).

Other less prevalent ICs identified in this study have been found in other countries. IC4 has been described in Europe and Brazil. CTX-M-115-carrying isolates from IC6 have been reported in Russia, Italy, and Germany (53).

cgMLST analysis allowed us to identify concrete clusters of isolates that share more than 99% of their allelic profiles in the country, thus establishing clonal groups present in the country’s different regions. The presence of ST79^P^ isolates, specifically from MST-1, which are multidrug-resistant, could be a public health problem due to its prevalence in patients with HAIs. Genomic surveillance and the identification of these patterns in *A. baumannii* can contribute to preventing the dissemination and propagation of resistance elements.

We found that *bla*OXA-23-like β-lactamases were the most frequent resistance element after the intrinsic chromosomal genes *bla*OXA-51-like and ADC-2 within the isolates. Interestingly, the isolates that did not present *bla*OXA-23-like β-lactamases carried *bla*OXA-24-like, *bla*OXA-143-like or other enzymes, such as KPC-2 or NDM-1. *bla*OXA-like β-lactamases are one of the most important determining factors that contribute to resistance to carbapenems, which has also been described in other studies in the country and the world (50,54).

The detection of the phenotypic coexistence of carbapenemases is considered a challenge (55). In the case of *Acinetobacter* spp., the detection of coproduction of carbapenemases using inhibitors can be difficult, considering that the most common carbapenemases are class D or OXAs and for these enzymes, there is no efficient inhibitor (56), and their confirmation requires molecular methods, such as polymerase chain reaction (PCR). In this study, nine coproductions were found with other carbapenemases. Coproduction of NDM-1 and *bla*OXA-23 was previously reported (57), and our results demonstrate the spreading of NDM-1 to other regions of the country with different genetic backgrounds. The *bla*NDM gene in *Acinetobacter* species is frequently detected worldwide, including in Latin America (58), and is mainly carried by the family of plasmids known as pNDM-BJ01-like (59). Carbapenemase VIM-4 was present as a coproduction in 3 isolates transported in plasmids. VIM-4 was first described in a *Pseudomonas aeruginosa* isolate from Greece, and *bla*VIM genes were carried on gene cassettes in class 1 integrons, which can disseminate rapidly (60,61). KPC-2 in *A. baumannii,* previously identified in Puerto Rico in Latin America, (62) was observed in one isolates, ST945^P^, carried on the transposon Tn4401 from the Santander department. This is the first report of this carbapenemase in *A. baumannii* in Colombia. The circulation and spread of carbapenemase-coproducing strains are of concern because some of these isolates may be associated with increased resistance to carbapenems and other antibiotics, making their treatment a challenge (63–65).

In A. *baumannii,* quinolone resistance is determined predominantly by alterations in the quinolone resistance-determining regions (QRDRs) of *gyrA* (Ser83Leu, Gly81Asp) and *parC* (Ser80Leu, Glu84Lys, Gly78Cys) (66). Following our results, 77 isolates (92.8%) presented the Ser83Leu and Ser80Leu mutations in *gyrA* and *parC*, respectively. Two new mutations in the *gyrA* (Ser83>Ala) and *parC* (Ser80>Ala) genes were identified in phenotypically resistant isolates. Additional analyses are recommended to confirm the role of resistance due to these new mutations. Aminoglycoside resistance in *A. baumannii* is mainly associated with modifying enzymes, such as acetyltransferases (AAC), phosphotransferases (APH), and adenyltransferases (ANT) (66). Our analysis showed that APH(3’’)-Ib and APH(6)-Id were the most frequently identified enzymes (86.7% and 84.3%, respectively).

The comparison of the phenotypic resistance profiles with the genetic elements identified in each isolate showed significant concordance. It allowed us to directly relate the resistance to each antibiotic with the elements found. The most common genomic resistance profile was associated with IC5 isolates resistant to all antibiotics. In general, in the 83 isolates, genes coding for the 4 resistance mechanisms were identified, a characteristic that makes *A. baumannii* multidrug-resistant. This condition helps it survive in an environment of pressure from antibiotics since the presence of several mechanisms makes it possible to avoid antibiotics in different ways.

This study was limited to isolates from voluntary surveillance at the national level, nevertheless, it is the study with the most departments included so far in the territory. The results offer a general overview at the genomic level of the isolates of *A. baumannii* that circulate in Colombia, and the information obtained is essential for national surveillance.

A strategy to prevent the appearance and dissemination of CRAB in health institutions is necessary to establish and strengthen policies related to the proper use of antibiotics, improve molecular surveillance, and adapt genomic surveillance to better monitor this pathogen.

## Ethical Approval Statement

This study did not involve human subjects, identifiable human data, or biological samples. The bacterial isolates were obtained through voluntary laboratory-based surveillance. Therefore, no IRB or ethics committee approval, patient consent, or clinical trial registration was required. All relevant ethical guidelines have been followed.

## Data Availability

All data produced in the present study are available upon reasonable request to the corresponding author. The genomic data and metadata used for analysis are referenced in the manuscript and supplementary materials.

https://microreact.org/project/cBr2FH4Mjge16gp7GopS64-acineto

## Acknowledgments

The authors thank the National Network of Laboratories for the isolates.

